# Excess Risk of COVID-19 to University Populations Resulting from In-Person Sporting Events

**DOI:** 10.1101/2020.09.27.20202499

**Authors:** Stephanie S. Johnson, Katelin C. Jackson, Matthew S. Mietchen, Samir Sbai, Elissa J. Schwartz, Eric T. Lofgren

**Affiliations:** Paul G. Allen School for Global Animal Health, Washington State University, Pullman, WA; Department of Mathematics and Statistics, Washington State University, Pullman, WA; School of Biological Sciences, Washington State University, Pullman, WA

## Abstract

**Background:** One of the consequences of COVID-19 has been the cancelation of in-person collegiate sporting events. We explore the impact of having in-person sports on COVID-19 transmission on a college campus, specifically the excess cases within the campus community can be anticipated.

**Methods:** Using a stochastic compartmental model representing the interactions between the university community, we model the impact of transient influxes of visitors attending sporting events and ancillary activities (bars, dining out, etc.). We consider a number of scenarios, varying the extent to which visitors interact with the campus, the number of infectious visitors, and the extent to which the campus has controlled COVID-19 absent events. We also conducted a sensitivity analysis, exploring the model’s outcomes over a wide range of uncertainty.

**Results:** Events caused an increase in the number of cases among the campus community, ranging from a 25% increase in a scenario where the campus already had an uncontrolled COVID-19 outbreak and visitors had a low prevalence of COVID-19 and mixed lightly with the campus community to an 822% increase where the campus had controlled their COVID-19 outbreak and visitors had both a high prevalence of COVID-19 and mixed heavily with the campus community. The model was insensitive to parameter uncertainty, save for the duration a symptomatic individual was infectious.

**Conclusion:** In-person sporting events represent a threat to the health of the campus community. This is the case even in circumstances where COVID-19 seems controlled both on-campus and among the larger population visitors are drawn from.

## Introduction

Enmeshed in the ongoing debate about the safety and necessity of in-person, on-campus university classes during the COVID-19 pandemic is a debate about whether collegiate level in-person sporting events can safely be held. This debate is most notably – though not exclusively – centered around collegiate football, whose regular-season takes place in the fall, and whose worth to the colleges that host these games is measured in the tens if not hundreds of millions of dollars. Additionally, these events involve a large number of players and coaching staff, and frequently draw a large crowd of spectators, potentially numbering in the tens of thousands of individuals. At present, several athletic conferences, including the Ivy League, Big Ten, and Pac-12, among others, have canceled or delayed their seasons already. However, as universities reopen and attempt to salvage their football season, these decisions are likely to be revisited.

There are many facets to the question of whether or not in-person sporting events can be rendered safe, ranging from how to prevent transmission between players during the game (which has been demonstrated for other pathogens^1,2^), how to enforce safe social distancing practices within the stadium, and how players, coaches and support staff may safely gather to practice and travel to games. Addressing all of these questions is beyond the scope of any one study. In many cases, it requires intensive setting specific knowledge ranging from the epidemiology of COVID-19 to the built environment of locker rooms. In this study, we attempt to address one specific COVID-19 related question, which can be applied more generally to many collegiate sports settings, especially those in rural areas: what is the impact of periodic, short-duration influxes of individuals from outside the immediate surrounding community who are there specifically to attend an in-person sporting event as well as engage in ancillary activities, such as tailgating, socializing at bars and restaurants, and otherwise potentially mixing with an otherwise somewhat isolated student and resident community? To address this, we use a mathematical model to simulate just such a periodic influx, considering the impact on the health of the student population.

## Methods

### Transmission Model

The transmission of COVID-19 between university students and from visiting spectators to students was modeled as a series of compartments representing various population health states (Figure 1). Students were divided into five health states – Susceptible (capable of being infected – S), Exposed (infected but in the latent period and not yet infectious – E), Infected (infected and symptomatic – I), Asymptomatic (infected but have mild or no symptoms – A) and Removed (Having recovered from their infection and having immunity for the remainder of the simulated period – R). Visitors from outside the campus community were modeled as being either Susceptible (V_S_) or Transmitting (V_T_) – this status is set at the beginning of the simulation and is assumed to remain static, representing an overall population of visitors at a given prevalence level, rather than the specific infection course of individuals.

**Figure 1.**
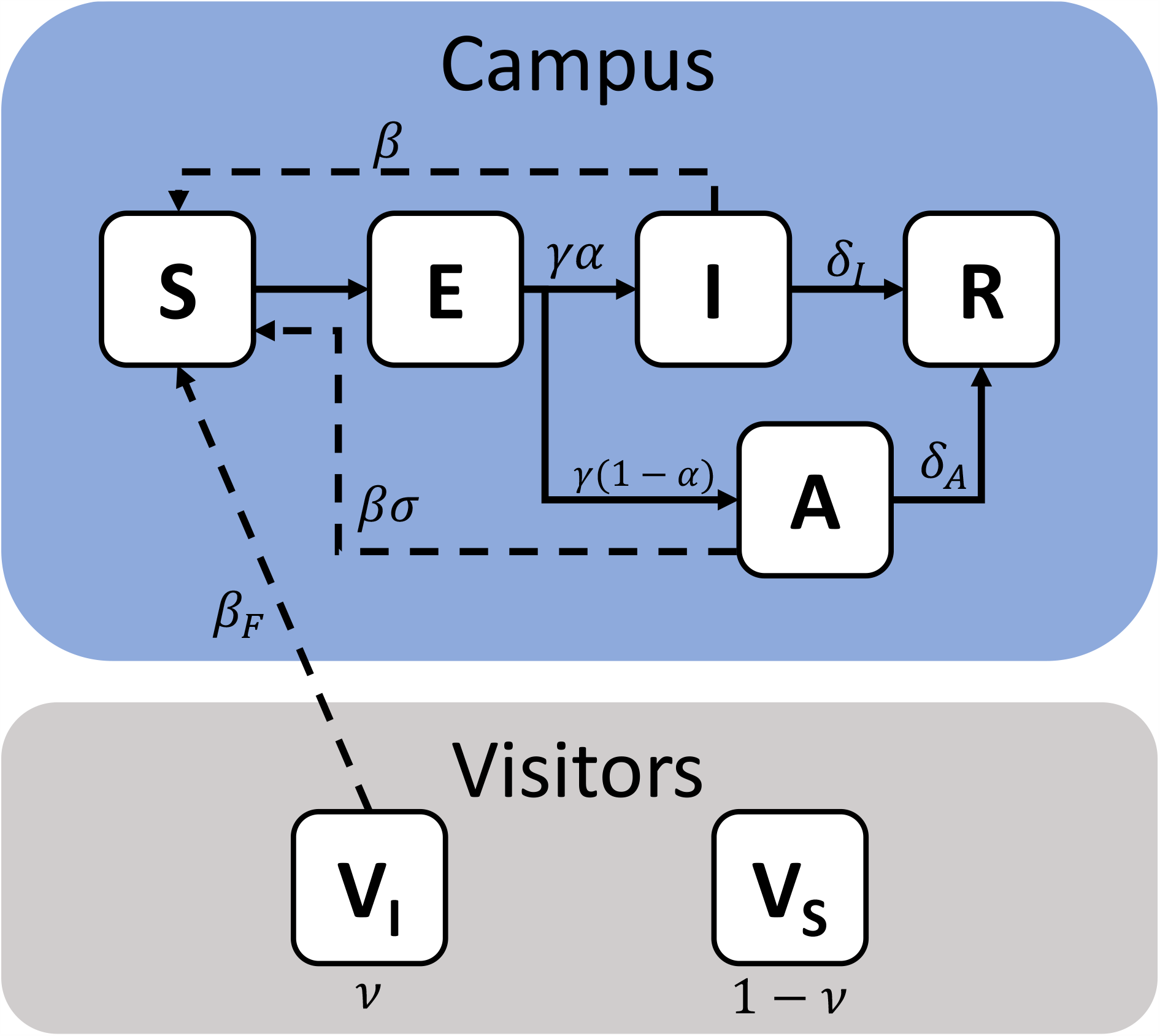
Schematic representation of a model of COVID-19 transmission due to in-person sporting events. The campus population is shown within the blue field, while visitors are shown within the grey field. The campus population can take four health states – Susceptible (S), Exposed (E), Infectious (I), Asymptomatic (A) or Recovered (R), while visitors are represented as two states, Susceptible (V_S_) or Infectious (V_I_). Solid arrows denote transition terms, while dashed arrows denote potential transmission pathways. The relevant parameters for each are also shown.

The interactions between these compartments was governed by a series of eight stochastic transitions, the details of which are presented in Table 1. The model was parameterized from the literature, with the value and meaning of the parameters, as well as their sources, similarly reported in Table 2. The recorded outcome was the number of incident infections among students during a 109-day period, representing the beginning of the school year up until the beginning of winter break. This model makes several simplifying assumptions – it is assumed that all students are equally exposed to visitors and otherwise mix randomly with each other. In order to simplify the model, and due to the very low rate of severe symptoms or death in this age group, we do not separately model hospitalizations or deaths. Rather, these are projected out from the number of incident infections that occur in the model (i.e. transitions from S to E). Finally, the model does not attempt to fully track the epidemic taking place outside the student population, instead focusing on the group for which the hypothetical modeled university has the greatest responsibility to safeguard their health and well-being.

**Table 1.** Transitions and equations for a model of COVID-19 transmission due to in-person sporting events

| Transition | Equation |
| --- | --- |
| S → E | $\beta S \frac{I}{C} + \beta \sigma S \frac{A}{C} + \beta_F S \frac{V_I}{C + V}$ |
| E → I | $\gamma \alpha E$ |
| E → A | $\gamma (1 - \alpha) E$ |
| I → R | $\delta_I I$ |
| A → R | $\delta_A A$ |
| C | S + E + A + I + R |
| V | $V_S + V_I$ |

**Table 2.**
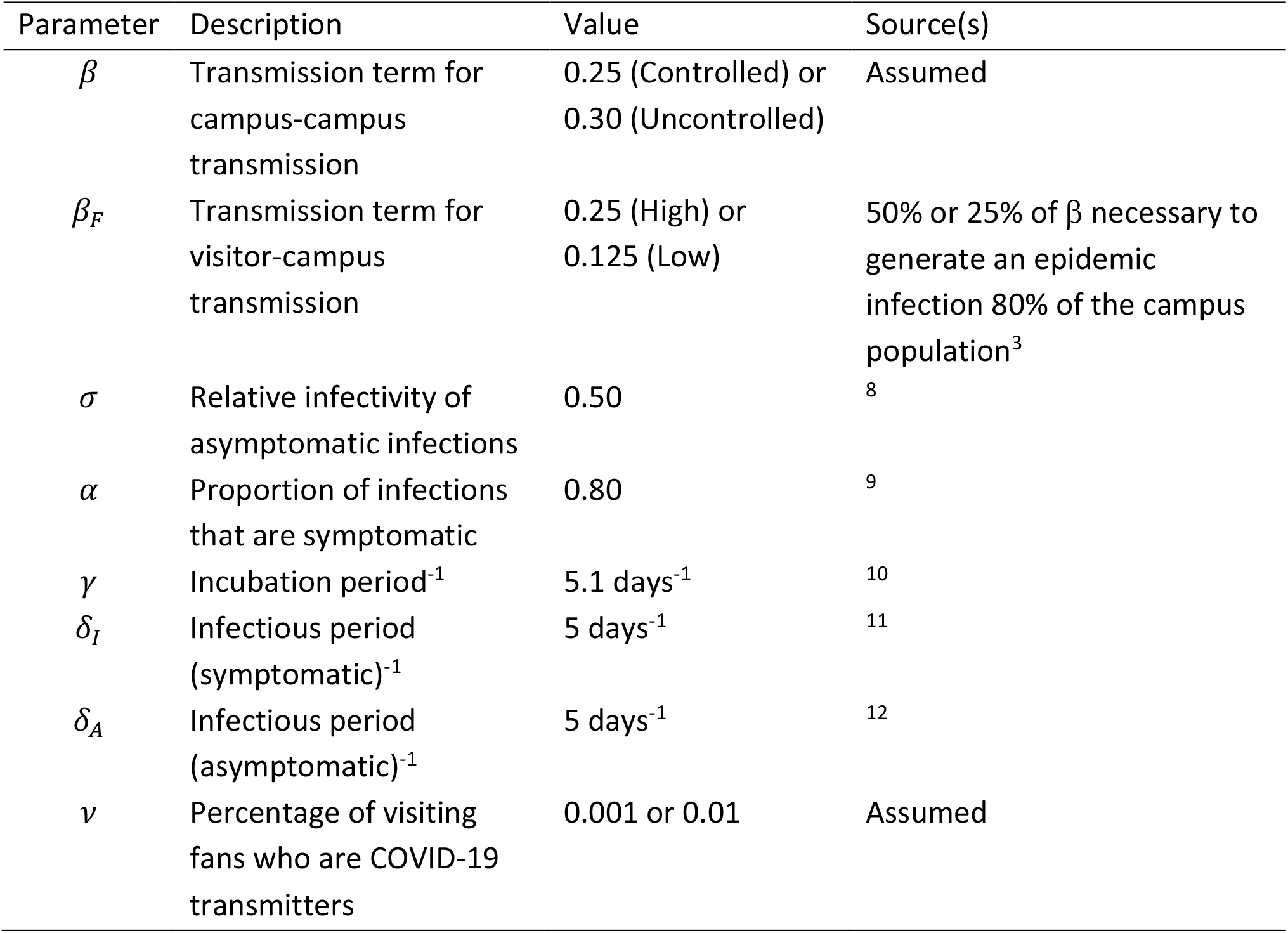
Parameters for a model of COVID-19 transmission due to in-person sporting events

| Parameter | Description | Value | Source(s) |
| --- | --- | --- | --- |
| $\beta$ | Transmission term for campus-campus transmission | 0.25 (Controlled) or 0.30 (Uncontrolled) | Assumed |
| $\beta_F$ | Transmission term for visitor-campus transmission | 0.25 (High) or 0.125 (Low) | 50% or 25% of $\beta$ necessary to generate an epidemic infection 80% of the campus population<sup>3</sup> |
| $\sigma$ | Relative infectivity of asymptomatic infections | 0.50 | <sup>8</sup> |
| $\alpha$ | Proportion of infections that are symptomatic | 0.80 | <sup>9</sup> |
| $\gamma$ | Incubation period<sup>-1</sup> | 5.1 days<sup>-1</sup> | <sup>10</sup> |
| $\delta_I$ | Infectious period (symptomatic)<sup>-1</sup> | 5 days<sup>-1</sup> | <sup>11</sup> |
| $\delta_A$ | Infectious period (asymptomatic)<sup>-1</sup> | 5 days<sup>-1</sup> | <sup>12</sup> |
| $\nu$ | Percentage of visiting fans who are COVID-19 transmitters | 0.001 or 0.01 | Assumed |

### Modeling Existing Campus-wide COVID-19 Control

Because the effectiveness of on-campus control of COVID-19 remains unknown, we considered two scenarios to represent the campus community’s attempts at mask wearing, social distancing, and other COVID-19 related interventions expressed as two differing values of the transmission term between students, *β*. In the first case, we consider a scenario where the community has, by and large, successfully implemented COVID-19 controls, driving the basic reproductive number (R_0_) slightly below one. Small, stochastically driven and largely self-limiting outbreaks are possible, but the unchecked transmission is highly unlikely. The second scenario has a slightly higher value for *β*, representing a community may have successfully “flattened the curve” and slowed the epidemic, but has not been able to lower R_0_ sufficiently to prevent a major outbreak over the course of the Fall semester. These scenarios were selected to represent universities that could, credibly, still be attempting to have students (including student-athletes) on campus. Higher values of *β* representing uncontrolled outbreaks were not represented, as it seems likely in these scenarios that a university- or state-level decision to close the campus would, by necessity, also impact in-person sporting events.

### Representing In-Person Sporting Events

In-person sporting events (hereafter called “events”) were modeled based on a series of event triggers within the simulation. When events are not taking place, the interaction between visitors and students is assumed to be zero. When an event is triggered, for two days, visitors interact with the student population, governed by the parameter *β*_*F*_. This represents the mixing not only taking place during the sporting event itself, but also ancillary activities such as tailgating, patronizing the same bars and restaurants, etc. The events were triggered on Days 18, 25, 39, 46, 67, and 88 of the simulated 109-day semester, using the at-home scheduled football games for Washington State University as a prototypical schedule (https://wsucougars.com/sports/football/schedule/2020).

### Modeled Scenarios

We considered a total of eight scenarios, representing a range of possible circumstances in which the decision to hold or cancel events might be made. For each of the two community control scenarios (i.e. R_0_ > 1 or R_0_ <1), we modeled the prevalence of COVID-19 infective individuals within the visiting population at 0.01 and 0.001, dubbed “High Prevalence” and “Low Prevalence” respectively. These represent visitors coming from areas with largely uncontrolled COVID-19 transmission or areas with significant but controlled COVID-19 transmission.

In combination with this, we modeled two different scenarios for how the student and visiting populations interact during the game itself as well as ancillary activities such as parties, tailgating, visiting bars and restaurants, etc. The first (“High Mixing”) assumed that the student-visitor interaction rate (*β*_*F*_) was 50% of the student-student interaction rate, while the second (“Low Mixing”) assumed the student-visitor interaction rate was 25% of the student-student interaction rate. Importantly, as we believed that either students or visitors engaging in these activities were inherently less likely to be following other social distancing guidelines, they were modeled off the student-student interaction rate for an *uncontrolled* epidemic. As such, the value for *β*_*F*_ was derived by multiplying a value of *β* that produced an epidemic infecting 80% of the student population (a common early estimate for the impact of an uncontrolled COVID-19 outbreak^3^) by 0.5 or 0.25, respectively.

Each scenario was stochastically simulated for 1000 iterations using Gillespie’s Direct Method^4^ in Python 3.8 using the using the StochPy library^5^. Statistical comparisons between scenarios were performed using a non-parametric Kruskal-Wallis test, with pairwise-comparisons performed via Pairwise Wilcoxon Rank Sum Tests, adjusting for multiple comparisons using the Benjamini and Hochberg method^6^.

### Sensitivity Analysis

We conducted a semi-global sensitivity analysis to quantify the impact of uncertainty in other parameters in addition to the main scenario-based analyses. In this analysis, we chose to examine the sensitivity of a potential policy-relevant outcome – the relative increase in cases between having and not having in-person events. To do this, we chose the scenario set with the most pronounced internal difference: a campus that had achieved an R_0_ < 1.0 without events vs. that same campus with events using the High Prevalence and High Mixing Scenarios. The five free parameters in that model (*α,δ*_I,_ *δ*_A_, *γ* and *σ*) were allowed to vary uniformly ± 50% of the value used in the main analysis (−50% to +25% in the case of *α* to ensure, as a probability, that its value did not exceed 1.0). A random set of these varied parameters was drawn, and then simulated 25 times with and without events. The median of the 25 simulations with events was then divided by the median of the 25 simulations without to provide an estimate of the relative increase in COVID-19 cases due to events. This process was repeated 1,000 times, and linear regression was used to estimate the relative impact of a single percentage change in each parameter value on this estimated relative burden due to events.

Additionally, to explore the impact of the relatively speculative values used for how intensely the visiting and student populations mix compared to pre-COVID-19 campus-campus interactions (25% or 50% in all the scenarios above), we conducted an additional sensitivity analysis focusing on *β*_*F*_, varying this from 0 to 0.50, representing a range of possibilities from equivalent to having no in-person events to having the same mixing rate as the campus community would have with itself in the pre-COVID-19 era. Ranges above this are theoretically possible but were not considered. A value for *β*_*F*_ was drawn from a uniform distribution between 0 and 0.50, and a single epidemic was simulated with this value. To thoroughly explore the parameter space and variability of outcomes, this process was repeated 10,000 times. The parameter draws were then rescaled to be a percentage of relative mixing 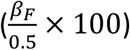 for ease of interpretability, and a Poisson regression model was fit to estimate the number of cases arising from a one-unit increase in this measure. In this scenario, we assumed the other parameters remained fixed, and used the campus R_0_ < 1.0 and low COVID-19 prevalence in visitors as the baseline scenario.

### Software and Data Availability

All simulations were performed in Python 3.8 using the StochPy library^5^ while all statistical analysis was performed in R version 4.0. The code and simulation results are available at http://www.github.com/epimodels/inperson_sports.

## Results

### Baseline Models

The two control scenarios – where the control of COVID-19 on campus had achieved an R_0_ slightly below or slightly above 1.0 had a median of 216.5 cases in the student population versus 1118.5 respectively (*p* < 0.001), typifying largely small, stochastically driven outbreaks vs. uncontrolled epidemic spread (Figure 2).

**Figure 2.**
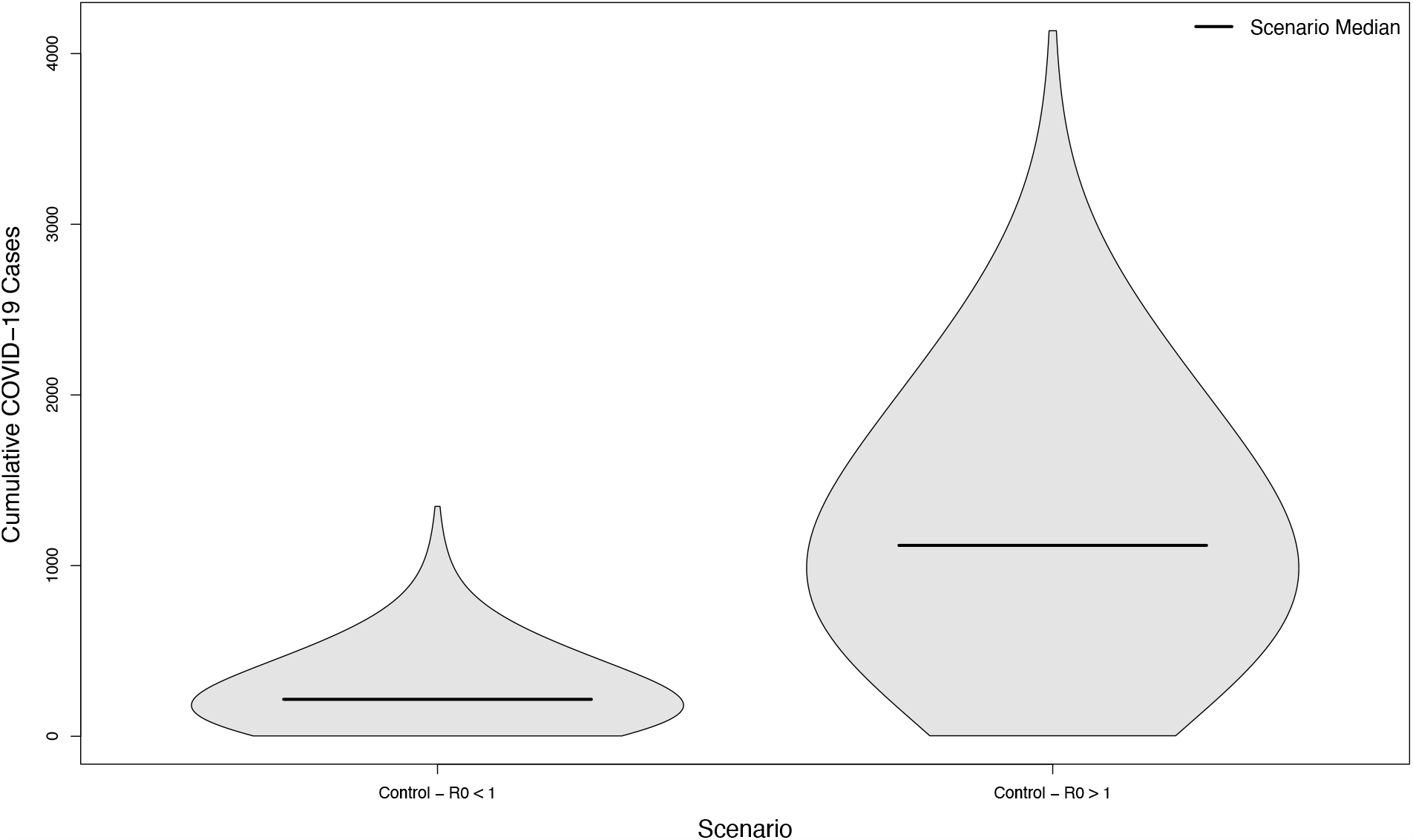
Distribution of cumulative COVID-19 cases in 1,000 simulated university campuses with no in-person sporting events, with and without control of their respective epidemics.

### Campus R_0_ < 1 Scenarios

Scenarios with and without in-person sporting events showed marked differences, even in scenarios where the overall campus R_0_ had been successfully reduced below one. Compared with the no event scenario’s median of 216.5 cases, scenarios with visitors from an overall low prevalence population had a median of 330 cases (+52%) assuming a low mixing rate between visitors and college students, and a median of 437 cases (+102%) assuming a high mixing rate between visitors and college students (*p* < 0.001 in both cases). The distribution of these simulated outcomes is shown in Figure 3. In scenarios with visitors from an overall high prevalence population, the median number of cases was 1196 (+452%) and 1997 (+822%), respectively. Both scenarios were statistically significantly different from their low prevalence counterparts (*p* < 0.001). The distribution of these simulated outcomes is shown in Figure 4.

**Figure 3.**
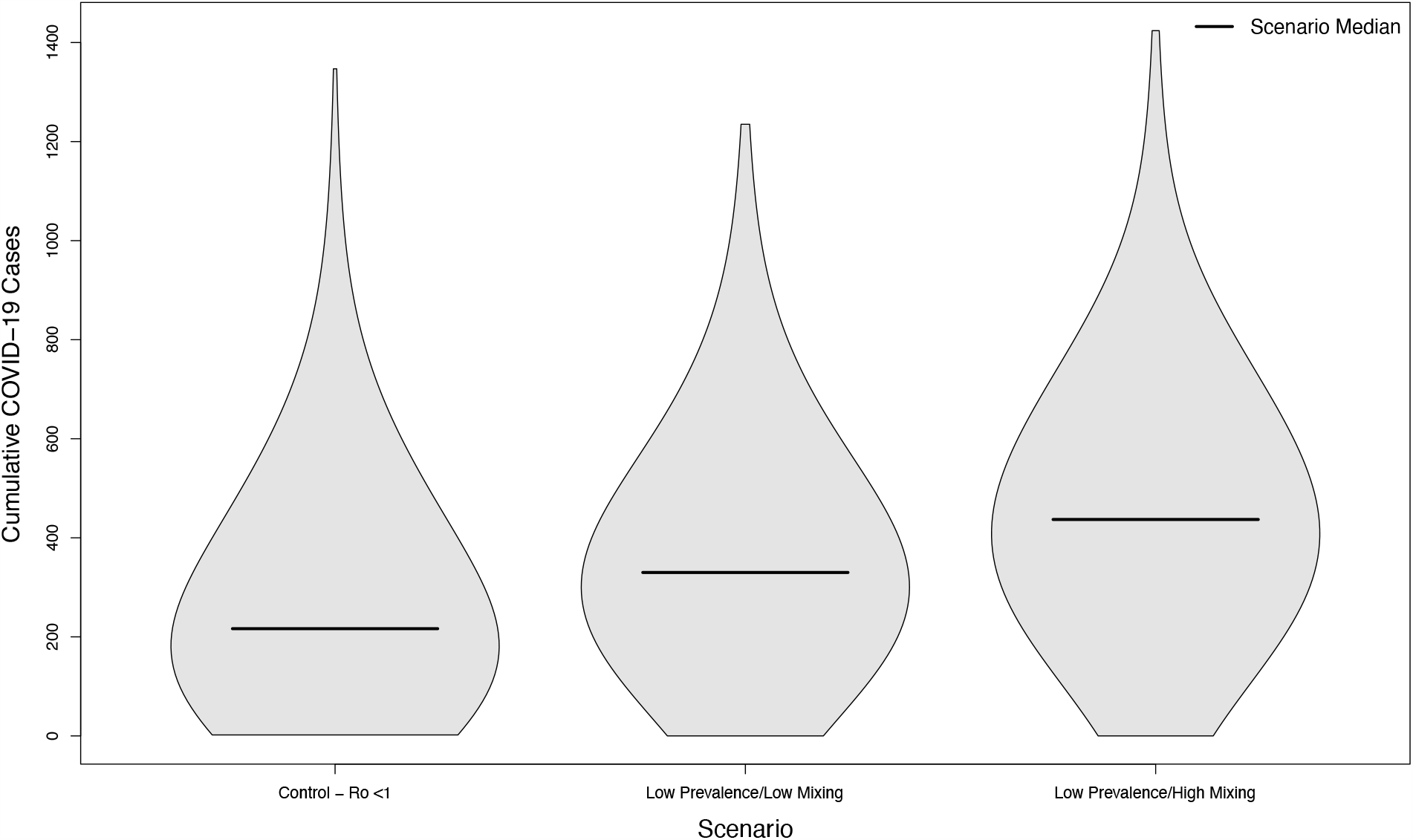
Distribution of cumulative COVID-19 cases in 1,000 simulated university campuses with controlled epidemics, comparing a campus with no in-person sporting events to two scenarios where visitors come from a low prevalence area and mix with the campus community at a low rate or a high rate respectively.

**Figure 4.**
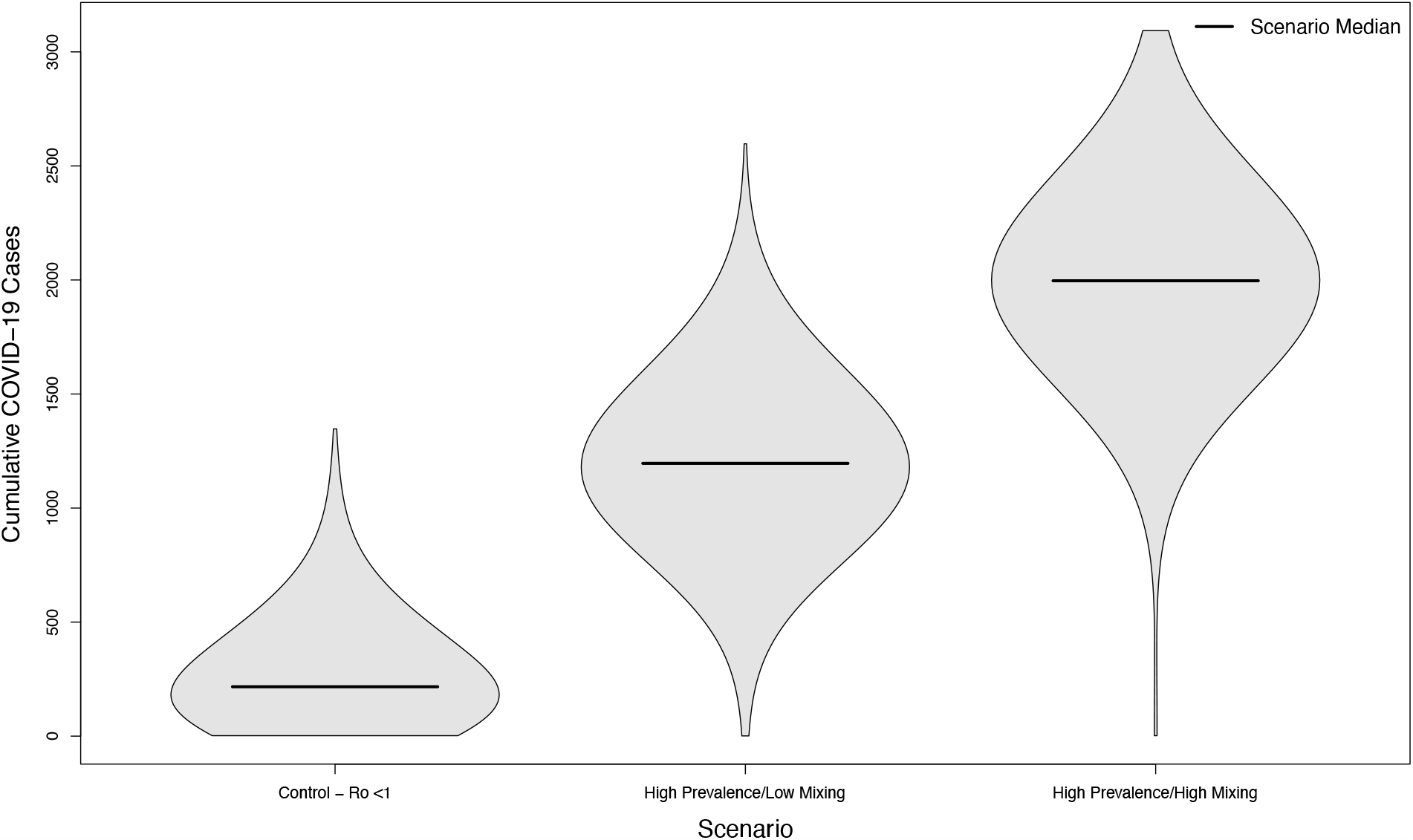
Distribution of cumulative COVID-19 cases in 1,000 simulated university campuses with controlled epidemics, comparing a campus with no in-person sporting events to two scenarios where visitors come from a high prevalence area and mix with the campus community at a low rate or a high rate respectively.

### Campus R_0_ > 1 Scenarios

Even in scenarios where the overall campus epidemic has not been successfully controlled, holding in-person sporting events increases the overall burden of disease. Compared to a baseline of 1118.5 cases without in-person sports, scenarios with visitors from an overall low prevalence population had a median of 1402 cases (+25%) assuming a low mixing rate between visitors and college students, and a median of 1596 cases (+43%) assuming a high mixing rate between visitors and college students (p < 0.001 in both cases). The distribution of these simulated outcomes is shown in Figure 5. In scenarios where visitors came from populations with an overall high prevalence, the median number of cases was 3102 (+177%) and 4522 (+304%) respectively. As previously, both scenarios were statistically significant from their low prevalence counterparts (*p* < 0.001). The distribution of these simulated outcomes is shown in Figure 6.

**Figure 5.**
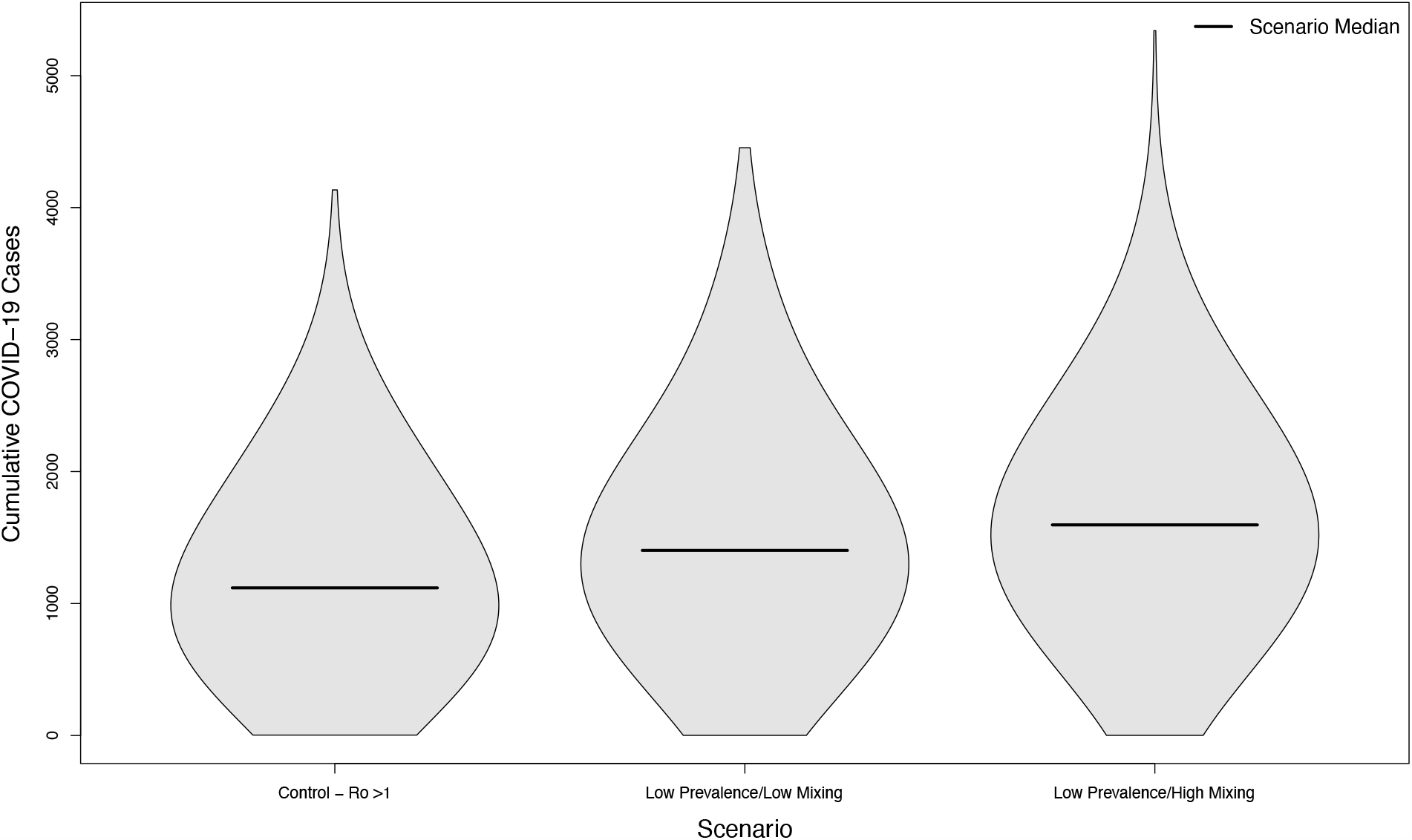
Distribution of cumulative COVID-19 cases in 1,000 simulated university campuses with uncontrolled epidemics, comparing a campus with no in-person sporting events to two scenarios where visitors come from a low prevalence area and mix with the campus community at a low rate or a high rate respectively.

**Figure 6.**
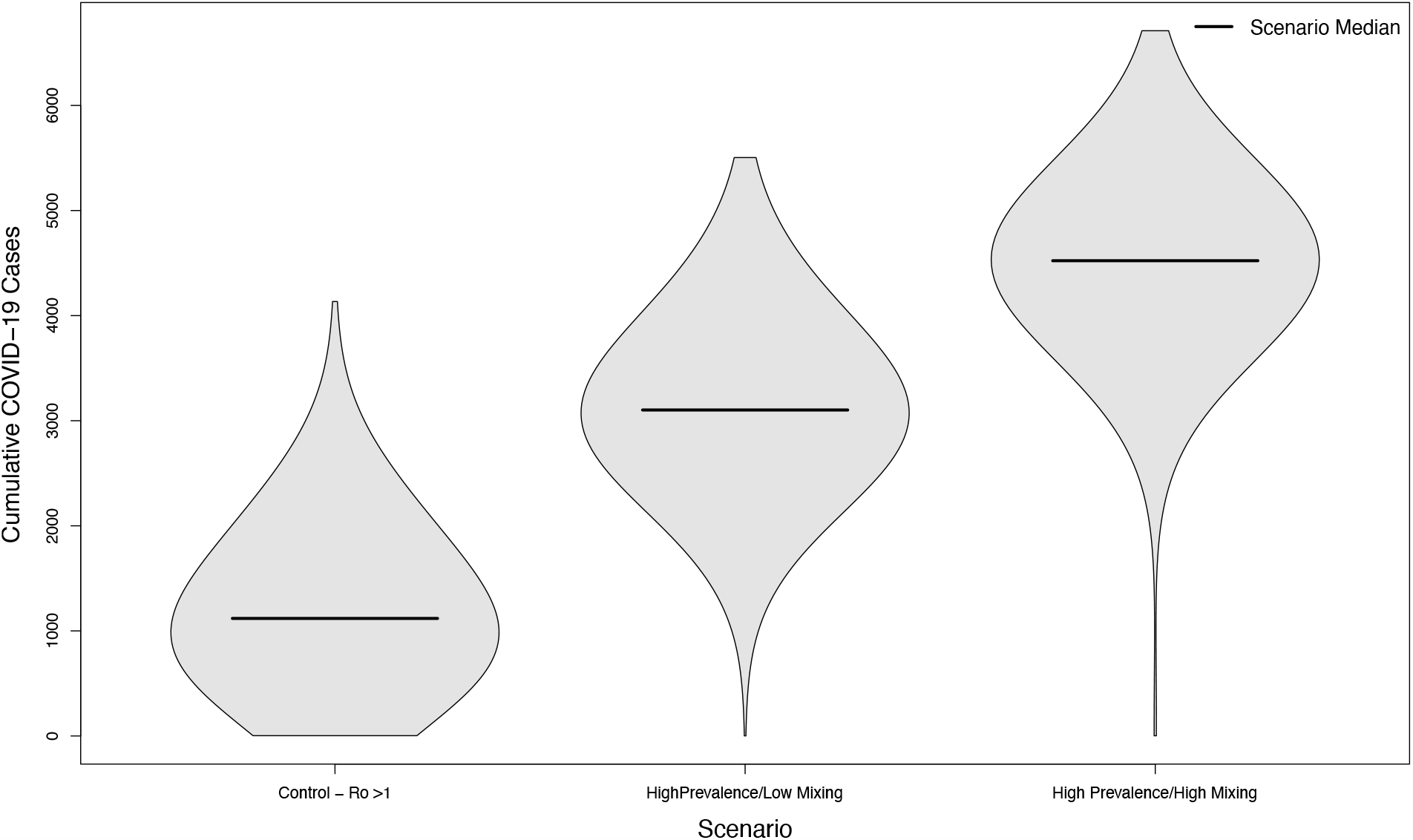
Distribution of cumulative COVID-19 cases in 1,000 simulated university campuses with uncontrolled epidemics, comparing a campus with no in-person sporting events to two scenarios where visitors come from a high prevalence area and mix with the campus community at a low rate or a high rate respectively.

### Sensitivity Analysis

The ratio of cases of COVID-19 between the scenario with no in-person sporting events vs. a scenario with sporting events drawn from a high prevalence setting and with high amounts of mixing between visitors and the campus community, where in both cases the campus R_0_ was below one, had a median of 11.08 (Interquartile Range: 4.78, 19.98). In a semi-global sensitivity analysis (*β* and ν are fixed) the model was relatively insensitive to changes in parameter values. A ten percent increase in a given parameter value results in a change of less than 0.6 in ratio for all but one parameter. This parameter was *δ*_*I*_—the duration during which an asymptomatic individual is infectious. In this case, a ten percent increase in *γ* (implying a shorter duration of infectiousness) increases the ratio by 2.75 (95% Confidence Interval 2.65 to 2.85). The sensitivity of the model to parameter changes is shown in Figure 7.

**Figure 7.**
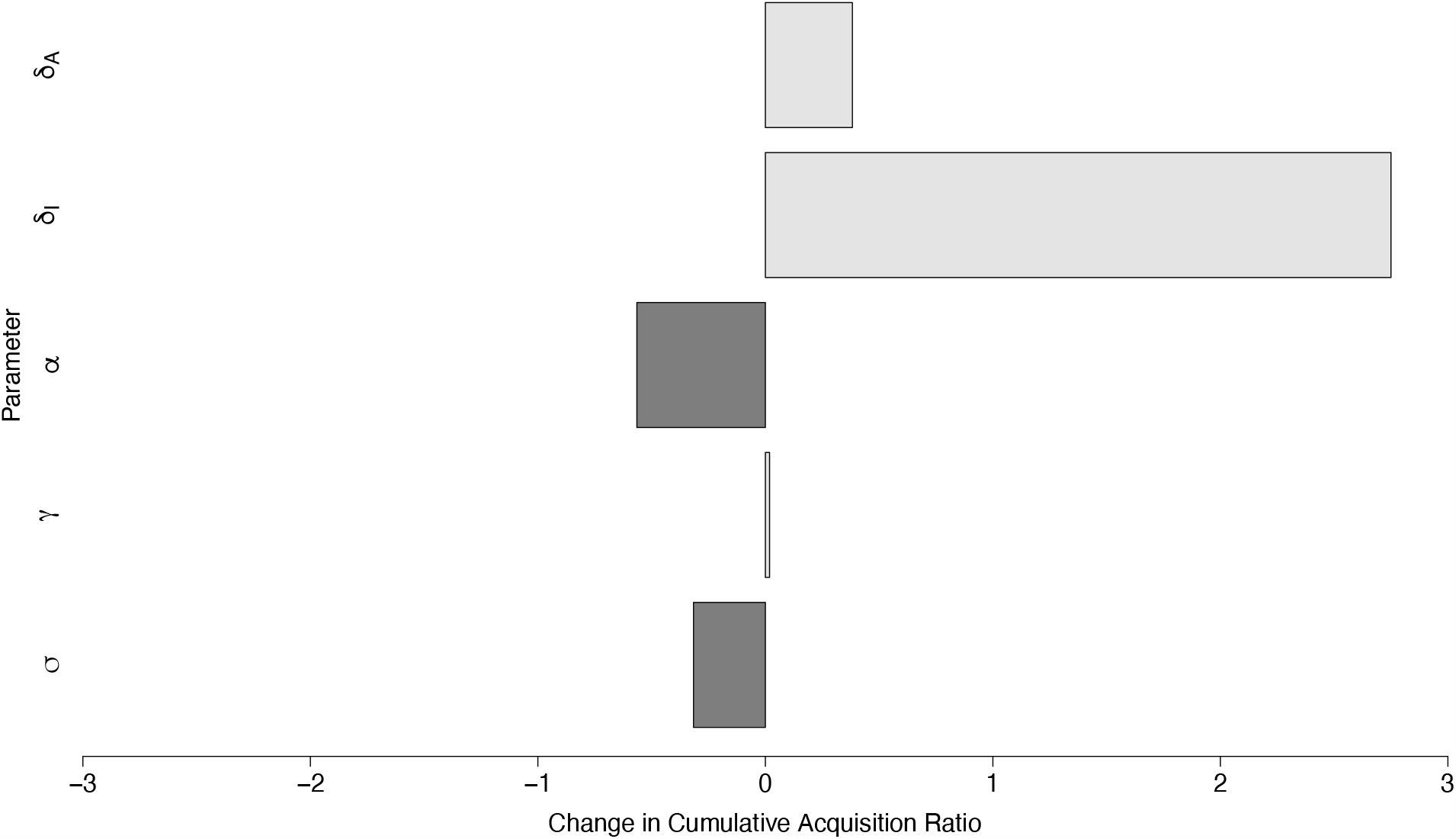
Global parameter sensitivity of the ratio of cases between two campuses with controlled epidemics – one with no in-person sporting events, and one where visitors come from a high prevalence area and mix with the campus community at a high rate. Each bar depicts the change in this ratio per ten-percent change in the value of a specific parameter.

The sensitivity of the model to *β*_*F*_ was as expected, with larger values corresponding to a higher number of cases (Figure 8). The Poisson regression model with a single term fit the relationship between the relative mixing rate between the community and visitors well, with a one-unit change in this rate (corresponding to an increase in *β*_*F*_ of 0.05) causing a relative increase in cases of 1.01 (95% CI: 1.01 to 1.01).

**Figure 8.**
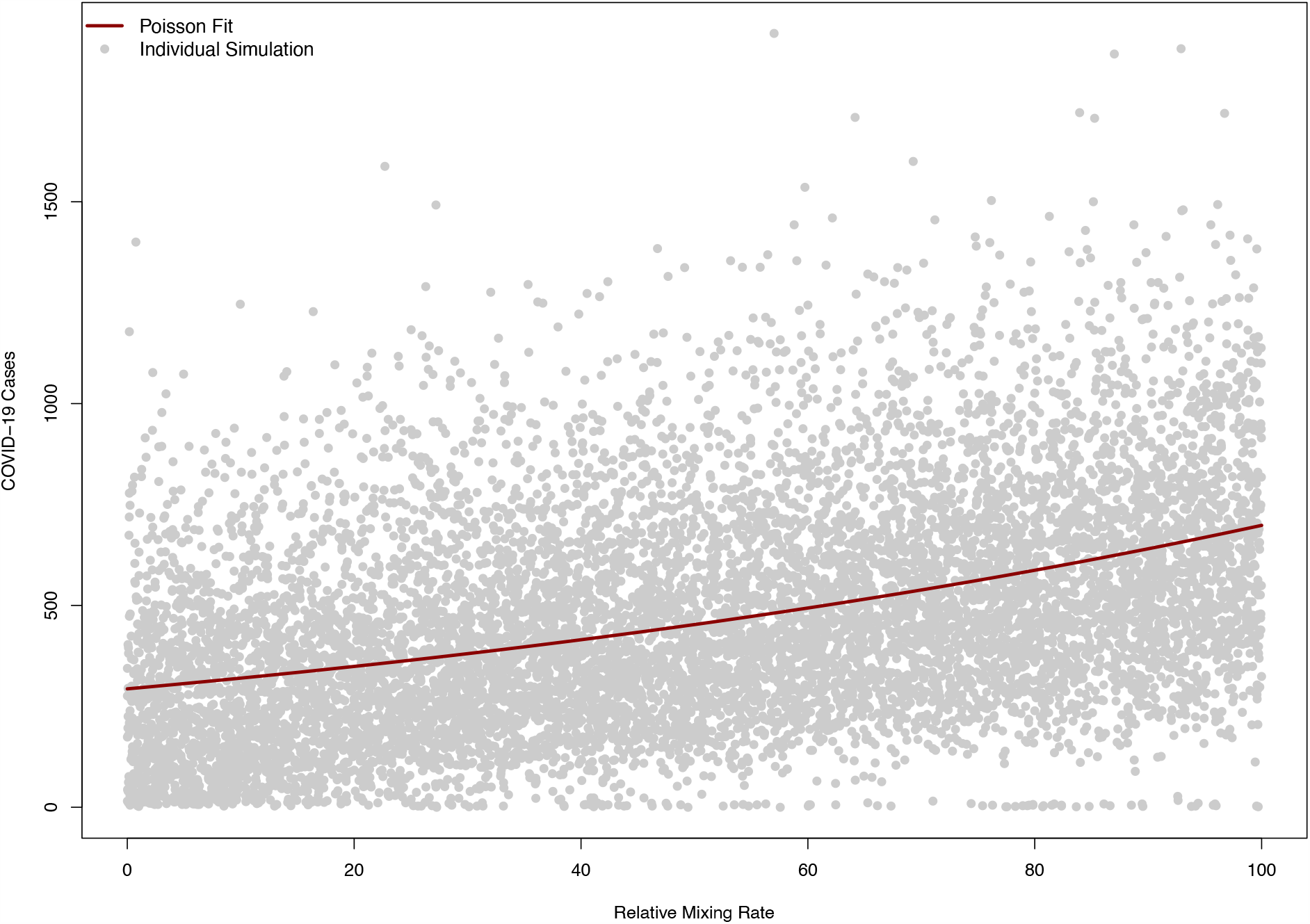
Model sensitivity to the relative mixing rate between campus community and visitors in a model (*β*_*F*_). A value of zero corresponds to no mixing between the campus and visitors, while 100 corresponds to a level of mixing equal to the campus-campus mixing rate pre-COVID. The solid red line indicates a Poisson regression fit of simulated COVID-19 cases, while each grey dot corresponds to a single simulation.

## Discussion

In all cases, allowing in-person sporting events to take place during a period of active COVID-19 transmission resulted in a significant increase in the overall burden of disease in the campus population. This impact was proportionately greater in campuses that otherwise had achieved reasonable levels of control over their epidemics, as the periodic bolus of seed cases stemming from in-person event attendance either re-ignited the epidemic or interrupted the descent of the epidemic toward stochastic extinction. In less controlled scenarios, although the proportional increase was smaller, in-person sporting events caused a larger increase in the absolute number of cases. In many circumstances, this may represent a more dangerous scenario, as a healthcare system’s capacity to absorb new cases and the corresponding adverse outcomes from COVID-19 is largely a question of fixed capacities (at least in the short term) of available beds, ventilators, etc., many of which are already constrained^7^.

The prevalence of COVID-19 in the source population of visitors had a large impact on the median number of cases in the simulation. Importantly however, even scenarios with only 10 infectious visitors out of a population of 10,000 were sufficient to cause substantial outbreaks. This means that risks to the campus community will persist even in areas where the overall rate of COVID-19 is relatively low. The results of the model are likely to be fairly robust to parameter uncertainty, and the modest, linear increase in cases caused by increasing values of *β*_*F*_ suggest that, while the “high mixing” and “low mixing” parameter values were somewhat speculative that the results of the model are not contingent on these specific values, and will scale readily if other values are likely in a given setting.

## Conclusion

The results of these simulations suggest that in-person sporting events represent a considerable health risk to the campus community. Even the most optimistic scenario modeled – visitors from an area with relatively low prevalence visiting a campus with a largely controlled outbreak and not mixing particularly heavily with the campus community, created 113 new COVID-19 student cases over the course of the semester. The ability to generate potentially hundreds of new cases even on both a controlled campus and in an area of low prevalence suggests that risk-free in-person sporting events are not necessarily possible for the foreseeable future.

More pessimistic scenarios were substantially worse, and the additional cases caused in these scenarios themselves represent serious COVID-19 outbreaks. It is also important to note that, in many senses, these scenarios are all optimistic – they do not represent direct transmission between players, in the community (i.e. transmission to wait staff, taxi or rideshare drivers, etc.) or between visitors themselves, but merely the cases that arise from visitor-student interactions. This was done because representing both the fine-scale, granular interactions that take place on the field and in the locker rooms, as well as the population-scale effects of transmission over the broad catchment area many teams draw from would be a modeling task of staggering complexity. The estimates here are, therefore, floors rather than ceilings in terms of the number of cases, and only express the numbers most directly relevant and measurable on a university campus. Additionally, it is important to note that in many circumstances, the conditions that cause these re-seeded epidemics to occur are beyond the control of the university itself. Students living off-campus, bars and restaurants serving both students and visitors during sporting events, parties, etc. may take place in ways that are difficult for university administrations to influence.

As with any modeling study, this study is not without limitations. Because the interactions between college students themselves and between college students and the surrounding community were poorly understood prior to COVID-19, several major assumptions must be made regarding those interactions. We assume random mixing (representing crowds at stadium choke points, local bars and restaurants, etc.) rather than more sophisticated mixing patterns that might be relevant for transmission. As noted previously, we also focus narrowly on the impact to the campus community as a necessary simplifying assumption. Finally, because the epidemiology of COVID-19 on college campuses is still in a nascent stage (albeit thus far one that suggests large outbreaks are the norm), this model does not reproduce any *specific* campus experience with in-person sporting events, but rather explores a set of hypothetical scenarios we never the less believe are reasonable. Finally, this model only estimates infections, rather than more serious outcomes or deaths. The model’s outcomes were restricted to cases only, as the rate of severe outcomes in the college-aged population is still not well understood, especially for long-term impacts from moderate or severe cases of COVID-19. As these statistics become better known, extending the results of these models to represent those worse outcomes is straightforward.

With these caveats in mind, we believe our results show that in-person sporting events represent a significant public health hazard even under conditions that may seem to suggest they can be conducted relatively safely, and are potential sources for a vast number of infections in circumstances where COVID-19 is not otherwise well controlled. Given the uncertainty about the seasonal dynamics of COVID-19, if any, and the current difficulties college campuses are facing as they reopen, it seems prudent from a public health perspective to seriously consider the cancelation of in-person sporting events on college campuses for the duration of the pandemic, or at least until a widespread and effective vaccine is available.

## Data Availability

The code and simulation results are available at http://www.github.com/epimodels/inperson_sports.

## Acknowledgments

SSJ, KCJ, MSM and ETL are supported by Contract 75D30120P07911 from the Centers of Disease Control and Prevention.

## Notes

### Competing Interest Statement

The authors have declared no competing interest.

### Author Declarations

This manuscript is a modeling study using existing published data, and as such does not require IRB approval.

